# Evidence of ongoing *Trypanosoma cruzi* transmission in a low-infestation area of eastern Bolivia

**DOI:** 10.1101/2025.07.10.25331250

**Authors:** Bratriz Amparo Rodríguez-Olguin, Daniel F. Lozano Beltrán, Isabel Mariscal Sejas, Brandon N. Mercado-Saavedra

## Abstract

Chagas disease remains a major public health concern in Latin America, with Bolivia reporting one of the highest burdens of infection. While congenital transmission has become the predominant route of new infections in several countries, vectorial transmission persists in rural and peri-urban regions. Postrervalle, in the department of Santa Cruz, is officially classified as a low-infestation area; however, updated community-level data remain limited.

**Methods:** We conducted a cross-sectional study in July 2023 involving 58 mothers and 104 of their children in Postrervalle. Participants underwent serological screening using three diagnostic assays, and epidemiological data were obtained through structured maternal questionnaires. Logistic regression models were used to assess associations between child *Trypanosoma cruzi* seropositivity and maternal or household exposures during pregnancy.

**Results:** Seroprevalence was 15.5% among mothers and 3.8% among children. Notably, all seropositive children were born to mothers who tested seronegative, suggesting non-congenital transmission. In multivariable analysis, living in houses with mud walls during pregnancy was strongly associated with child seropositivity (adjusted OR = 225.44), while older child age also increased the odds of infection. Other maternal exposure variables showed elevated but imprecise associations.

**Conclusion:** Despite its classification as a low-infestation area, Postrervalle shows evidence of ongoing transmission of *T. cruzi* linked to domestic structural conditions that facilitate triatomine colonization. These findings highlight the importance of integrating entomological surveillance with household-level risk assessments to better characterize and prevent Chagas transmission in rural communities.

**AUTHORS’ SUMMARY:** Chagas disease is caused by the parasite *Trypanosoma cruzi* and is mostly spread by insects known as “kissing bugs,” which can live in the cracks of walls and roofs in some homes. Although several Latin American countries have reduced transmission through long-term vector control programs, Chagas disease continues to affect many rural and peri-urban communities in Bolivia. Postrervalle, where this study was carried out, is officially classified as a low-risk area, and little recent information exists about local transmission patterns. We screened mothers and their children for Chagas disease and asked the mothers about their living conditions and past exposure to kissing bugs. We found that none of the infected children had mothers who were infected, which suggests that these children did not acquire the disease during pregnancy. Instead, they were likely infected after birth through contact with infected insects in their homes. We also found that children who lived in houses with mud walls were more likely to be infected, indicating that certain types of housing may make it easier for kissing bugs to enter or hide inside homes. These findings show that Chagas transmission can still occur in places thought to be low-risk. Strengthening vector surveillance and improving housing conditions could help reduce infections in similar communities.

## INTRODUCCION

Chagas disease (CD) is caused by the protozoan *Trypanosoma cruzi* and is prevalent in most countries of Latin America ^1–3^. It is estimated that there are currently over 6 million people infected with the parasite and more than 70 million people at risk of getting the infection ^1,4^. CD has different modes of transmission, including the vectorial, congenital, and by blood transfusion ^1,3^. It is estimated that 1 out of 3 people infected with the parasite might progress to Chagas Cardiomyopathy, one of the chronic manifestations of CD ^5–7^. Among these countries, Bolivia is particularly affected, presenting both the vector and congenital transmission, making it a critical context for the study of CD ^2,3,5^.

CD is a neglected infectious disease in several countries, including Bolivia, which is estimated to have the highest prevalence of the infection in the world ^8^. The high prevalence of CD in Bolivia might be due to the presence of the vector in several areas of its territory, occupying over 40% of the communities in the country, according to the Bolivian National Program of Chagas disease (BNPCD) ^9^. A research study evaluated the prevalence of CD in a rural community in the Gran Chaco, a region highly endemic for the vector in Bolivia, presenting a prevalence that ranged from 40-80% ^10^. A community-based study performed in Spain in people from Latin American countries described that 40% of their participants came from Bolivia, and 36% of them were seropositive for CD ^11^.

Recent studies estimate that the prevalence of CD is approximately 12% among pregnant women in South America as of 2020 ^12^. In Bolivia, data from the BNPCD reported a prevalence of 2.4% in children aged 5 to 15 years and a congenital transmission rate of 1.4% in 2019 ^13^. By 2022, the National Program of Vector-Borne Diseases (ETVs) estimated a chronic infection prevalence of 19.2% among individuals over 15 years of age ^14^. Although vector control initiatives and expansions in maternal and child health care have led to declines in severe morbidity and mortality associated with Chagas disease in Bolivia, evidence of a sustained reduction in vectorial transmission remains inconclusive ^12,13^. Subsequent surveillance data have not consistently confirmed the decreases reported in earlier program evaluations ^15^. A 2017 study conducted in Camiri, a municipality in the Gran Chaco region of Bolivia, confirmed a seroprevalence of 0.22% among children aged 5 to 21 years, with rates increasing with age and a mean age of seropositivity of 13 years ^16^. Similarly, studies in the urban areas of Cochabamba reported prevalence rates ranging from 19% to 25% among children aged 5 to 13 years, reinforcing that Chagas disease remains not only a rural but also an urban public health concern ^17^.

This study aims to describe the epidemiological characteristics of Chagas disease among mothers and their children in the community of Postrervalle, a locality officially classified as having a low vector infestation rate by the Bolivian National Chagas Program ^9^. By examining seroprevalence patterns and associated risk factors, this study aims to determine whether vector-borne transmission within the community is the primary source of infection among children.

## METHODS

### Study Population

This is a cross-sectional, non-probabilistic, exploratory study of mother–child pairs. The study population included 58 mothers with their children (104 in total), ranging from 4 to 17 years old. The recruitment took place in July 2023 in the Postrervalle municipality, located in the southwest of the department of Santa Cruz, Bolivia (Figure 1). According to the 2024 national census ^18^, Postrervalle has a population of 1,804 inhabitants. Of these, 528 are children and adolescents aged 0–17. The municipality has only two schools, one elementary school and one high school, with a combined enrollment of 342 students. The study population was based on the students from these two schools. All students with their mothers were invited to participate in the study. We enrolled a total of 57 mothers and 104 students (children); 17 mothers were recruited with one child, 27 mothers with two children, 10 mothers with 3 children, and 4 mothers with 4 children. Nine children refused to participate, despite their mother and siblings participating in the study. All mothers in the study had at least one child.

**Figure 1.**
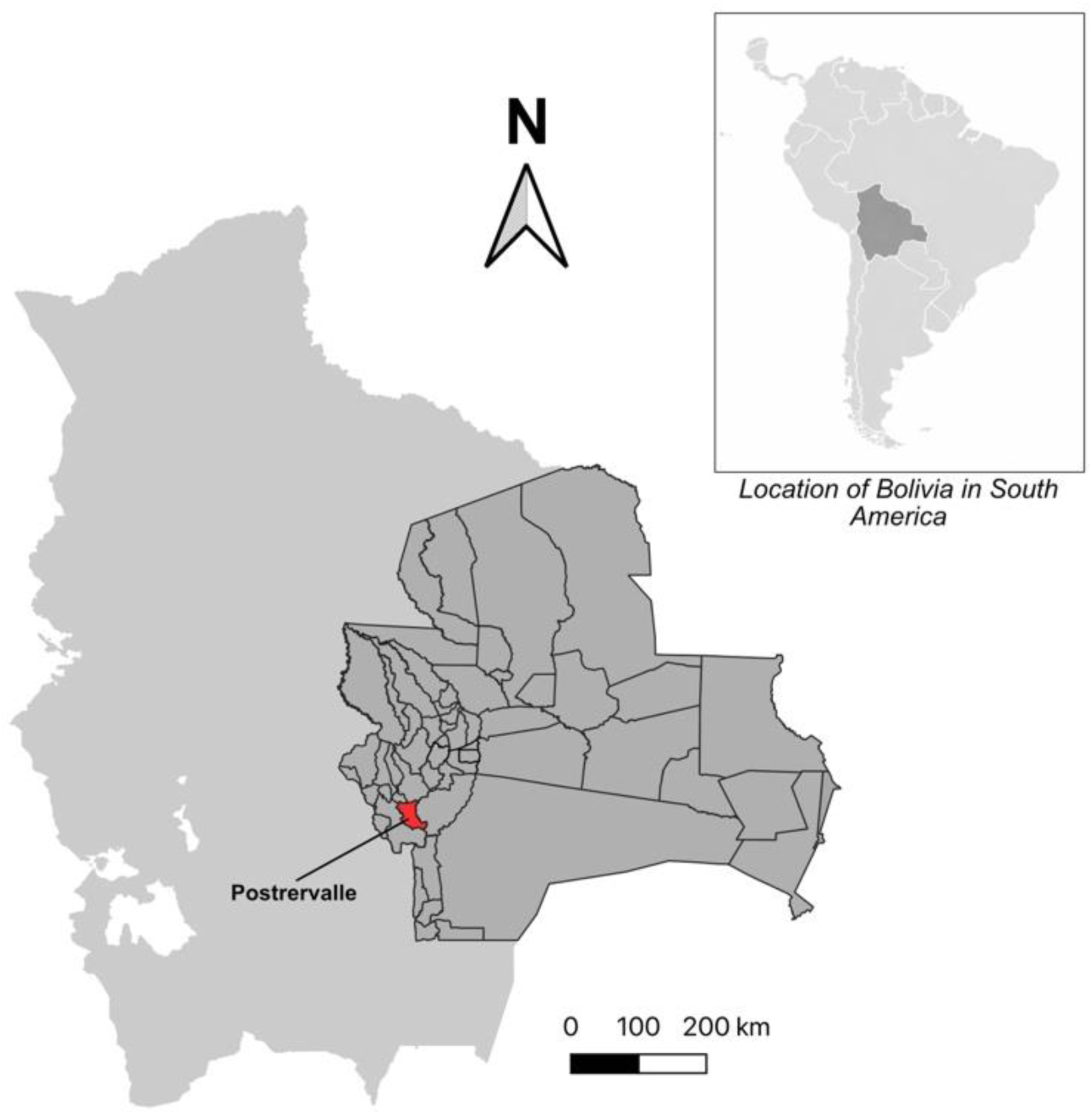
Location of Postrervalle in Santa Cruz, Bolivia. The upper right corner map shows the location of Bolivia in South America. The map of Bolivia in light gray shows the location of the department of Santa Cruz (dark gray) and its communities. Postrervalle is located at the southwest of Santa Cruz.

The recruitment was initiated by presenting the project to all the students’ parents at a meeting organized by the head of the school. Both the heads of the school and the parents’ representatives agreed that the project would take place in the schools in the following week. Mothers were asked to visit the schools on a certain date with their children to participate in the study.

All participating mothers completed an epidemiological questionnaire that included sociodemographic information, details about household characteristics (such as wall materials), and questions related to each of their pregnancies (including Chagas testing, history of blood transfusions, and presence of kissing bugs in the home). The pregnancy-related section was administered separately for each pregnancy reported by the mother

### Specimen collection and sample processing

Blood samples were collected from all consenting students and their mothers. The samples were allowed to coagulate and centrifuged at 3500 rpm for clot and serum separation. Serum was aliquoted and stored at -20°C. The serum was used to perform three different serological tests: OnSite Chagas Ab Combo Rapid Test—CTK BIOTECH (USA), Chagatest HAI—Wiener lab (Rosario, Argentina), and Chagatest ELISA recombinant v.3.0—Wiener lab (Rosario, Argentina). Participants who had at least two positive serological tests were considered Chagas seropositive. Participants with only one positive serological test were considered seronegative. All Chagas-positive participants (children and their mothers) were referred to the National Program of Chagas in the municipality for follow-up and treatment according to the national regulations.

### Statistical Analysis

The data were analyzed using Stata 18.0 (StataCorp, College Station, US). Continuous variables were summarized as means ± standard deviations (SD) or medians with interquartile ranges (IQR), depending on the normality of their distribution assessed by the Shapiro–Wilk test. Categorical variables were described as absolute frequencies and percentages. Comparisons between Chagas-seropositive and seronegative groups were conducted separately for mothers and children. For continuous variables, Student’s t-test was applied when normality and homoscedasticity assumptions were met, while the Mann–Whitney test was used for non-normally distributed data. For categorical variables, the Chi-square test or Fisher’s exact test (when expected cell counts were < 5) was used to assess associations between serological status and epidemiological factors.

The primary outcome was *T. cruzi* seropositivity, defined as reactivity in at least two of the three serological assays performed. Independent variables included demographic factors (e.g., maternal age, number of children), housing characteristics (wall, roof, and floor materials, fumigation history), and exposure-related variables (history of blood transfusion, presence of triatomine bugs, being bitten by kissing bugs, and previous Chagas testing or treatment). For children, maternal exposure variables were analyzed as potential risk factors for infection using a Firth’s penalized likelihood logistic regression adjusted and adjusted for sex and age.

All statistical tests were two-tailed, and a p-value < 0.05 was considered statistically significant. Missing data were minimal (< 5%) and excluded from specific analyses as pairwise deletions. Graphical representations (age distribution of seropositive vs. seronegative children) were produced using Stata’s graph box to visualize distributional differences.

### Ethics statement

The research protocol was revised and approved by the Institutional Review Board (IRB) of the Bolivian Catholic University in Santa Cruz, code 032, on 26/07/22. The recruitment was initiated on 24/07/2023 and finalized on 27/07/2023, following prior meetings with local authorities and school representatives that took place several months before the recruitment date. Each mother provided written informed consent for herself and her child. In addition, children provided assent to participate in the study. Seropositive participants were referred to the Chagas National Program for treatment and clinical follow-up in the medical center at Postrervalle.

## RESULTS

### Demographic characteristics of mothers

The study included 58 mothers, of whom 49 (84.5%) were seronegative for *T. cruzi*, and 9 (15.5%) were seropositive for *T. cruzi* (Table 1). The median age of the mothers was 37 years (IQR: 32 – 42), with no significant difference between seronegative and seropositive groups (p=0.494). Both seropositive and seronegative mothers had a median number of 2 children (p=0.463). Although 49 (86%) of mothers reported having been tested for Chagas disease at some point in their lives, the proportion was slightly higher among seropositive mothers (88.5%) – even though the difference was not significant (p=0.783). Nevertheless, the same number of mothers (4) from each group (seropositive and seronegative) reported being bitten by a kissing bug, the proportion was significantly higher in seropositive mothers 4 (44.4%) compared to seronegative mothers 4 (8.3%; p=0.004). Additionally, seropositive mothers were more likely to receive Chagas treatment (22.2% vs. 2.1%, p=0.014).

**Table 1.** Demographic and Exposure Characteristics of Mothers Stratified by Chagas Serology Status (N=58)

| Variables | Serology status for Chagas in mothers |  |  | p value |
| --- | --- | --- | --- | --- |
|  | Negative<br>49 (84.5%) | Positive<br>9 (15.5%) | Total<br>58 (100%) |  |
| Mother's age | 37 (32 – 41) | 38 (36 – 44) | 37 (32 – 42) | 0.494! |
| Number of children | 2 (1 – 3) | 2 (2 – 2) | 2 (1 – 2) | 0.463! |
| Chagas test (yes)* | 41 (85.4%) | 8 (88.9%) | 49 (86.0%) | 0.783 |
| Received blood transfusion (yes)* | 5 (10.4%) | 0 (0.0%) | 5 (8.8%) | 0.311 |
| Seen kissing bugs (yes)* | 7 (14.6%) | 3 (33.3%) | 10 (17.5%) | 0.175 |
| Lived in rural areas (yes)* | 18 (37.5%) | 1 (11.1%) | 19 (33.3%) | 0.123 |
| Been bitten by a kissing bug (yes)* | 4 (8.3%) | 4 (44.4%) | 8 (14.0%) | 0.004 |
| Received treatment for Chagas (yes)* | 1 (2.1%) | 2 (22.2%) | 3 (5.4%) | 0.014 |
| House received fumigation* | 31 (64.6%) | 8 (88.9%) | 39 (68.4%) | 0.150 |
| Mud wall (yes)* | 13 (27.1%) | 2 (22.2%) | 15 (26.3%) | 0.761 |

\* During any pregnancy
! Mann-Whitney test
All other tests were performed using $X^2$ Fisher's exact test
Numeric variables (mother's age and number of children) are represented by Median (IQR)

### Chagas Serology in Mothers and Children

Chagas serology testing was performed on 58 mother-child groups. Each mother was paired with all their children included in the study. As mentioned in Table 1, the prevalence of Chagas seropositivity among mothers was 15.5%. Table 2 describes that 6.9% of mothers had seropositive children. All seropositive children are born to seronegative mothers; in all cases, all three serological tests were negative for mothers and positive for their children.

**Table 2.** Chagas Serology Results of Mothers and Their Children (N=58)

|  | Mothers' Chagas serology |  | Total |
| --- | --- | --- | --- |
|  | Negative (%) | Positive (%) |  |
| **Children's Chagas serological status** |  |  |  |
| Negative | 45 (91.8%) | 9 (100.0%) | 54 (93.1%) |
| Positive | 4 (8.2%) | 0 (0.0%) | 4 (6.9%) |
| **Total** | 49 (84.5 %) | 9 (15.5%) | 58 (100%) |

Among children of seronegative mothers, 8.2% were seropositive, though this difference was not statistically significant.

### Factors Associated with Child *Trypanosoma cruzi* Seropositivity

A total of 104 children were included in the analysis, of whom 96.2% were seronegative, and 3.8% were seropositive for Chagas. Seropositive children were clustered at higher ages compared to seronegative children whose age was broadly distributed, although this difference was not significant (p=0.083) (Figure 2). Firth’s logistic regression analyses were conducted to identify factors associated with *T. cruzi* seropositivity among children, using epidemiological information reported by their mothers regarding exposures during pregnancy (Table 3). In the unadjusted models, child age was significantly associated with seropositivity (OR = 1.587; 95% CI: 1.052–2.395; *p* = 0.028), indicating that the likelihood of infection increased with age. No statistically significant associations were found with sex or being born in an urban setting.

**Figure 2.**
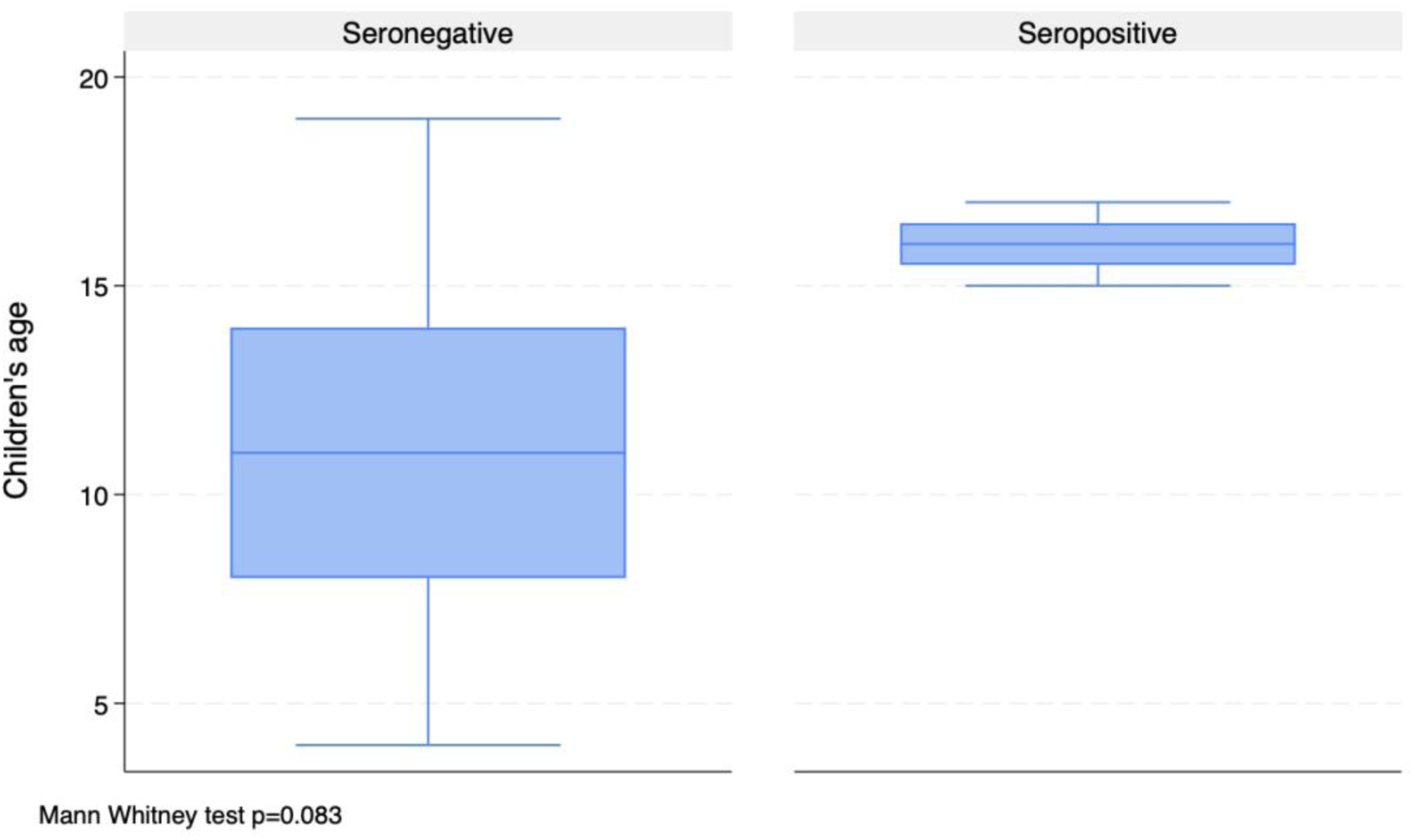
Age Distribution of Seropositive and Seronegative Children (N=104) Children’s age distribution by Chagas serostatus. The non-parametric Mann-Whitney’s p value is 0.083, suggesting a non-significant difference.

**Table 3.** Associated factors with *Trypanosoma cruzi* seropositivity among children, based on maternal and household exposures during pregnancy (N = 104).

| Variables | Unadjusted |  |  | Adjusted |  |  |
| --- | --- | --- | --- | --- | --- | --- |
|  | OR | [95% CI] | p value | OR | [95% CI] | p value |
| Age | 1.587 | [1.052 – 2.395] | 0.028 | 1.615 | [1.076 - 2.429] | 0.021 |
| Sex (women) | 1.433 | [0.237 – 8.650] | 0.694 | 2.779 | [0.376 - 20.525] | 0.316 |
| Lived in rural setting | 5.854 | [0.0.823 – 41.615] | 0.077 | 18.163 | [0.947 - 348.212] | 0.054 |
| Having a seropositive mother | 0.712 | [0.0362 - 13.981] | 0.823 | 0.079 | [0.001 - 6.239] | 0.255 |
| During pregnancy, mother |  |  |  |  |  |  |
| received blood transfusion | 7.285 | [0.0.895 - 59.327] | 0.063 | 37.858 | [0.736 - 1948.268] | 0.071 |
| saw kissing bugs in the house | 3.622 | [0.441 - 29.715] | 0.231 | 5.316 | [0.483 - 58.469] | 0.172 |
| was bitten by a kissing bug | 5.274 | [0.625 - 44.492] | 0.126 | 7.711 | [0.614 - 96.780] | 0.114 |
| received Chagas treatment | 28.428 | [2.329 - 346.893] | 0.009 | 10.864 | [0.639 - 184.793] | 0.099 |
| live in house with mud wall (yes) | 8.628 | [1.202 - 61.941] | 0.032 | 38.566 | [1.897 - 783.894] | 0.017 |

Firth's penalized likelihood logistic regression
Adjusted by age and sex

Among exposure variables reported during pregnancy, two maternal factors showed elevated odds in unadjusted analyses: previous Chagas treatment (OR = 28.428; 95% CI: 2.329–346.893; *p* = 0.009) and living in a house with mud walls (OR = 8.628; 95% CI: 1.202–61.941; *p* = 0.032). Although wide confidence intervals reflect small subgroup sizes, these findings suggest that domestic structural conditions and prior maternal infection history may be linked to child seropositivity.

After adjustment for child age and sex, the association between living in a house with mud walls and child seropositivity remained statistically significant (adjusted OR = 38.566; 95% CI: 1.897–783.894; *p* = 0.017). The relationships with maternal Chagas treatment, blood transfusion, and reported contact with kissing bugs showed elevated but non-significant adjusted odds ratios.

## DISCUSSION

Latin American countries have several neglected diseases, such as Chagas disease, with approximately 70 million people at risk of infection. Congenital transmission is the main source of infection in most countries in the region ^19,20^. Bolivia is a country with one of the highest burdens of Chagas disease, where multiple rural communities in the Chaco region are endemic to the vector ^8,9,21^. The official data presented by the BNPCD reports that Postrervalle is a low infestation community due to the low infestation rate of vectors (<1%)^14^. The BNPCH also reports that *Triatoma infestans* is the principal vector present in the region. In this study, we describe the situation of Chagas disease in mothers and children in Postrervalle, with a prevalence of mothers of 15.5%. This prevalence is similar to that reported by the local authorities, which estimate that the prevalence is 14.5% (official unpublished data). However, a recent meta-analysis described that the prevalence of pregnant women in Bolivia ranged from 17-65%, with lower rates in the past 5 years ^22^. This relates to a study that reports that the prevalence among pregnant women at the public maternity hospital in Santa Cruz is 22% ^23^. Nevertheless, these results were mainly hospital-based, with women from all regions (rural and urban) of the department, increasing the estimated prevalence of each community. One factor that is significantly more present among

Chagas seropositive mothers from this study was being bitten by the vector, where 4 (44%) of infected women reported having been bitten by a kissing bug. However, having lived in rural areas and living in houses with mud walls was not significantly more prevalent in women with Chagas disease. These results are not entirely consistent with other studies performed in similar settings ^10,11,24^, which might be due to the small sample size or the fact that most of the people included in the study have lived in Postrervalle for their entire lives, presenting similar epidemiological characteristics. One interesting finding was the case of a seronegative mother who had previously received treatment for Chagas disease. In this study, the mother tested negative across all three serological assays, which makes a current false-negative result unlikely. While treatment-induced seroconversion can occur, it is rare in chronically infected adults and typically requires many years of follow-up before antibody levels fall below detectable thresholds; it is most observed in infants or young children treated early in the course of infection ^25^. Therefore, given the mother’s adult age and the complete seronegative profile at the time of evaluation, the most plausible explanation is that she likely received a false-positive diagnosis in the past, rather than having undergone treatment-induced loss of detectable antibodies. These findings highlight the importance of contextualizing local seroprevalence patterns within broader regional transmission dynamics and historical vector control efforts.

Bolivia joined the Southern Cone Initiative, a 1991 collaborative program with Argentina, Brazil, Chile, Paraguay, and Uruguay, which aimed at interrupting Chagas transmission by eliminating *Triatoma infestans* and ensuring safe blood supplies ^26^. While countries like Uruguay, Chile, and Brazil successfully halted *Triatoma infestans* transmission by the late 1990s and early 2000s, Bolivia achieved interruption in parts of La Paz and Potosí only by around 2011–2013, though persistent bug populations and insecticide resistance continue to challenge complete eradication in regions such as the Gran Chaco ^27^. Notably, in our study, none of the mothers who were seropositive for *T. cruzi* had seropositive children, whereas all seropositive children were born to mothers who tested negative. In all cases, the three serological tests were consistently negative for the mothers and positive for the children, making false-negative or false-positive results unlikely. Because congenital transmission occurs only from infected mothers, this pattern indicates that infections in the children of our study are unlikely to be vertically acquired. Instead, these findings are more consistent with ongoing vectorial exposure in the domestic environment, a pattern also documented in childhood serosurveys in the Bolivian Chaco ^16,28^.

In Postrervalle, congenital transmission may be minimal, and instead, vectorial transmission is the likely source of infection among children. Although maternal screening was conducted at the time of this study, it is possible that some mothers may have seroconverted after giving birth or were false negatives due to limitations in the sensitivity of serological tests during acute or early chronic stages of infection. Alternatively, postnatal vector exposure is a plausible explanation, as all seropositive children were older than 8 years. This epidemiological pattern aligns with other studies conducted in rural or peri-urban areas of Bolivia with similar vector infestation classifications. For instance, Hopkins et al. (2019) reported persistent *T. cruzi* transmission in school-aged children in the Bolivian Chaco, even in areas under surveillance, with infection rates increasing with age and suggesting ongoing vector exposure in domestic settings ^16^. In another low-endemic setting in Cochabamba, Medrano-Mercado et al. (2008) found seroprevalence rates in children as high as 25%, reinforcing that low infestation classifications may not fully reflect the true risk of transmission, particularly in settings with structural vulnerabilities such as adobe walls and a lack of sustained vector control ^17^. Our logistic regression findings (Table 3) further support the role of domestic environmental conditions in transmission. Age was significantly associated with *T. cruzi* seropositivity among children. All seropositive cases occurred in children aged 15 years or older, while seronegative children showed a broader age distribution, suggesting cumulative exposure with increasing age. Most notably, living in a house with mud walls during pregnancy remained significantly associated with seropositivity after adjustment, indicating that household construction type may serve as a persistent ecological risk factor enabling vector colonization. Although other exposure-related variables (such as maternal history of kissing bug contact or prior Chagas treatment) showed elevated odds ratios, their wide confidence intervals likely reflect the small number of seropositive children and should be interpreted cautiously. Additionally, the potential emergence of pyrethroid resistance in *Triatoma infestans*, the principal domestic vector in Bolivia, is a growing concern. Resistance has been documented across several regions of the Gran Chaco, reducing the long-term effectiveness of indoor residual spraying campaigns ^29^. If similar resistance is present in Postrervalle, even a low measured infestation prevalence may mask ongoing domestic colonization and transmission. This reinforces the need to integrate routine entomological surveillance, community-based vector monitoring, and repeated serological screening of children—not only classifications of infestation levels—to accurately evaluate transmission risk. Finally, the characteristics associated with seropositivity in children included whether their mothers had seen or been bitten by kissing bugs, and whether they had lived in houses with mud walls, environments that likely reflect the domestic setting in which the children were born and spent their early childhood. These conditions represent key risk factors that facilitate vectorial transmission of *T. cruzi* ^23,30,31^. Taken together, these findings highlight that domestic ecological conditions continue to play a central role in sustaining vectorial transmission, even in communities labeled as low-risk.

This study has some limitations, including a relatively small sample size, the use of non-probabilistic sampling, and the fact that not all children of the participating mothers were included. Participant recruitment was conducted primarily through the only two local schools, where all the mothers were actively engaged. Although mothers whose children did not attend the school may have been underrepresented, it is unlikely that this introduced a significant selection bias, as there are no known sociodemographic or epidemiological differences between mothers with school-attending children and those whose children do not attend. This study included approximately 30% of registered students and 20% of all children in Postrervalle according to the latest national census, with an overall statistical power above 80%. Finally, not all children of the 58 participating mothers were enrolled, primarily due to the absence of verbal consent. Although this may influence the observed prevalence of Chagas disease in children, it is unlikely to have significantly impacted the general epidemiological patterns, as non-participating children are presumed to share similar risk profiles with their siblings.

These findings highlight that, even in areas officially classified as low infestation zones, vectorial transmission of *T. cruzi* persists as a route of infection among children. While Bolivia has made substantial progress under the Southern Cone Initiative, particularly in selected regions, the continued presence of infected vectors and active transmission in communities like Postrervalle underscores the fragility of these gains. Moreover, the overuse of insecticides—potentially leading to vector resistance—and the broader ecological impacts of climate change may further hinder vector control efforts, threatening to reverse recent advances and sustain transmission in vulnerable rural populations.

## Data Availability

All relevant data are within the manuscript and its Supporting Information files.

## AUTHORS’ CONTRIBUTION

BARO contributed to the conceptualization, study design, manuscript drafting, and coordination of fieldwork. DFLB was responsible for the methodological adaptation and refinement of the study design. IMS coordinated the sample collection team and supervised field procedures. BNMS conducted the data analysis, interpreted the statistical findings, and manuscript drafting.

## ACKNOWLEDGEMENTS

To Drs. Freddy A. Tinajeros and Nayrha Villazón, Lizeth Gil, and Mayra Morales for their contribution during different steps of this project. To Drs Benjamin Quiroga, Célida Montaño Gutiérrez, and Judith Aguilar from the Chagas Program of Santa Cruz for their support during this project. We also want to thank the community of Postrervalle who received us, and all the participants of the study.

